# Empowering Radiologists with ChatGPT-4o: Comparative Evaluation of Large Language Models and Radiologists in Cardiac Cases

**DOI:** 10.1101/2024.06.25.24309247

**Authors:** Turay Cesur, Yasin Celal Gunes, Eren Camur, Mustafa Dağlı

## Abstract

**Purpose:** This study evaluated the diagnostic accuracy and differential diagnosis capabilities of 12 Large Language Models (LLMs), one cardiac radiologist, and three general radiologists in cardiac radiology. The impact of ChatGPT-4o assistance on radiologist performance was also investigated.

**Materials and Methods:** We collected publicly available 80 “Cardiac Case of the Month’’ from the Society of Thoracic Radiology website. LLMs and Radiologist-III were provided with text-based information, whereas other radiologists visually assessed the cases with and without ChatGPT-4o assistance. Diagnostic accuracy and differential diagnosis scores (DDx Score) were analyzed using the chi-square, Kruskal-Wallis, Wilcoxon, McNemar, and Mann-Whitney U tests.

**Results:** The unassisted diagnostic accuracy of the cardiac radiologist was 72.5%, General Radiologist-I was 53.8%, and General Radiologist-II was 51.3%. With ChatGPT-4o, the accuracy improved to 78.8%, 70.0%, and 63.8%, respectively. The improvements for General Radiologists-I and II were statistically significant (P≤0.006). All radiologists’ DDx scores improved significantly with ChatGPT-4o assistance (P≤0.05). Remarkably, Radiologist-I’s GPT-4o-assisted diagnostic accuracy and DDx Score were not significantly different from the Cardiac Radiologist’s unassisted performance (P>0.05).

Among the LLMs, Claude 3.5 Sonnet and Claude 3 Opus had the highest accuracy (81.3%), followed by Claude 3 Sonnet (70.0%). Regarding the DDx Score, Claude 3 Opus outperformed all models and Radiologist-III (P<0.05). The accuracy of the general radiologist-III significantly improved from 48.8% to 63.8% with GPT4o-assistance (P<0.001).

**Conclusion:** ChatGPT-4o may enhance the diagnostic performance of general radiologists for cardiac imaging, suggesting its potential as a valuable diagnostic support tool. Further research is required to assess its clinical integration.

---

The advent of Large Language Models (LLMs) has marked a major advancement in artificial intelligence (AI). These models are trained on large datasets, allowing them to generate text that is both accurate and human-like (1). This technological breakthrough offers great promise for medicine and serves as a supportive tool in clinical decision making (1,2).

AI is becoming increasingly common in cardiac radiology. It is used for calcium scoring, evaluating coronary artery stenosis, analyzing plaque morphology, and assessing cardiac function and tissue characterization in cardiac MRI (3–6). The number of radiological examinations is increasing daily, but there are insufficient radiologists to interpret these scans. AI-assisted diagnostic systems are expected to reduce the workload of radiologists significantly (3).

LLMs have demonstrated outstanding performance in various radiological areas. They excel in evaluating radiological knowledge in board-style examinations, summarizing radiology reports for patients, and providing patient information (7–10).

The field of publicly available radiology cases, which focuses on patients’ medical histories and imaging findings, is a key area of research for comparing the performance of LLMs (11–17). Few studies have assessed the cardiac radiology knowledge of LLMs (17–19). Monroe et al. evaluated the performance of ChatGPT 3.5 and 4 on hypothetical cardiac imaging questions from a patient’s perspective (18). Sarangi et al. investigated the concordance and acceptance of five hypothetical cardiac imaging patterns generated by these models (19). Although these studies effectively addressed patients’ inquiries about cardiac radiology reports and theoretically demonstrated success in recognizing cardiac radiologic patterns, they did not provide results that accurately reflected everyday radiology practice. This is due to the absence of real-life patient history and radiological scenarios.

Li et al.’s study demonstrates the ability of LLMs to diagnose and generate differential diagnoses in cardiac radiology similar to real-life scenarios. However, in their study examining 287 “*Radiology* Diagnosis Please” cases, the performance of ChatGPT 3.5 and 4 was evaluated in only 17 cardiovascular cases (17). Additionally, only one study investigated ChatGPT-4 as an adjunct tool in general radiology (20). This study found that ChatGPT-4 contributes slightly to diagnostic accuracy and may partially mitigate the experience gap among radiologists (20).

As they also noted, the previously mentioned studies on the performance of LLMs in cardiac radiology have certain limitations (17,20). These include the narrow scope of question types covering different areas of cardiac radiology and the small number of questions and LLMs used in the studies (17,20). Additionally, to the best of our knowledge, no study has focused on the performance differences between ChatGPT-4o assisted and unassisted radiologists in cardiac radiology.

This study aims to evaluate the diagnostic accuracy and ability to generate differential diagnoses of 11 different LLMs (ChatGPT-4o, ChatGPT-4, ChatGPT-3.5, Google Gemini 1.5 Pro, Google Gemini 1.5 Flash, Google Gemini 1.0, Mistral Large, Meta Llama 3 70b, Perplexity, Claude 3 Opus, Claude 3.5 Sonnet and Claude 3 Sonnet), a cardiac radiologist, and three general radiologists using publicly available “Cardiac Cases of the Month” published on the Society of Thoracic Radiology website. We also aimed to investigate whether the performance of cardiac and general radiologists was affected by the assistance of ChatGPT-4o in the same set of cases.

## MATERIALS AND METHODS

### Study Design

This cross-sectional observational study evaluated the performance of various LLMs, including ChatGPT-4o, ChatGPT-4, ChatGPT-3.5, Google Gemini 1.5 Pro, Google Gemini 1.5 Flash, Google Gemini 1.0, Mistral Large, Meta Llama 3 70b, Perplexity, Claude 3 Opus, Claude 3.5 Sonnet and Claude 3 Sonnet. It also assessed the responses of three board-certified (EDiR) general radiologists and one cardiac radiologist when solving publicly available cardiac radiology cases. Ethics committee approval was not required as the study utilized only publicly available online cases. The study adhered to the Standards for Reporting Diagnostic Accuracy Studies (STARD) statement and principles of the Declaration of Helsinki (21).

### Data Collection

The Society of Thoracic Radiology has published monthly the “Cardiac Case of the Month” on its website (https://thoracicrad.org). We reviewed all 91 cases between December 2014 and September 2023. Seven cases were excluded because the answers were already provided, two were excluded because of duplicate entries, and two were excluded because they contained pathological information, resulting in a total of 80 cases for analysis.

These cases include a home page, Hint, Additional Images, Findings, Diagnosis, Discussion, and References sections. Both text-based and visual information exist within these sections. On the homepage, both the patient’s history (text-based information) and the first radiological image (visual information) were presented. The Additional Images section contains supplementary radiological images of the patient as visual information. In the Findings section, the patient’s radiological findings are provided as text-based information.

The Diagnosis section includes the patient’s diagnosis and differential diagnosis information. To better showcase the performance of radiologists and LLMs in our study, we did not use the Hint section as a source of information. Figure 1 illustrates the layout of the anonymized homepage for a typical case and the aforementioned sections.

**Figure 1.**
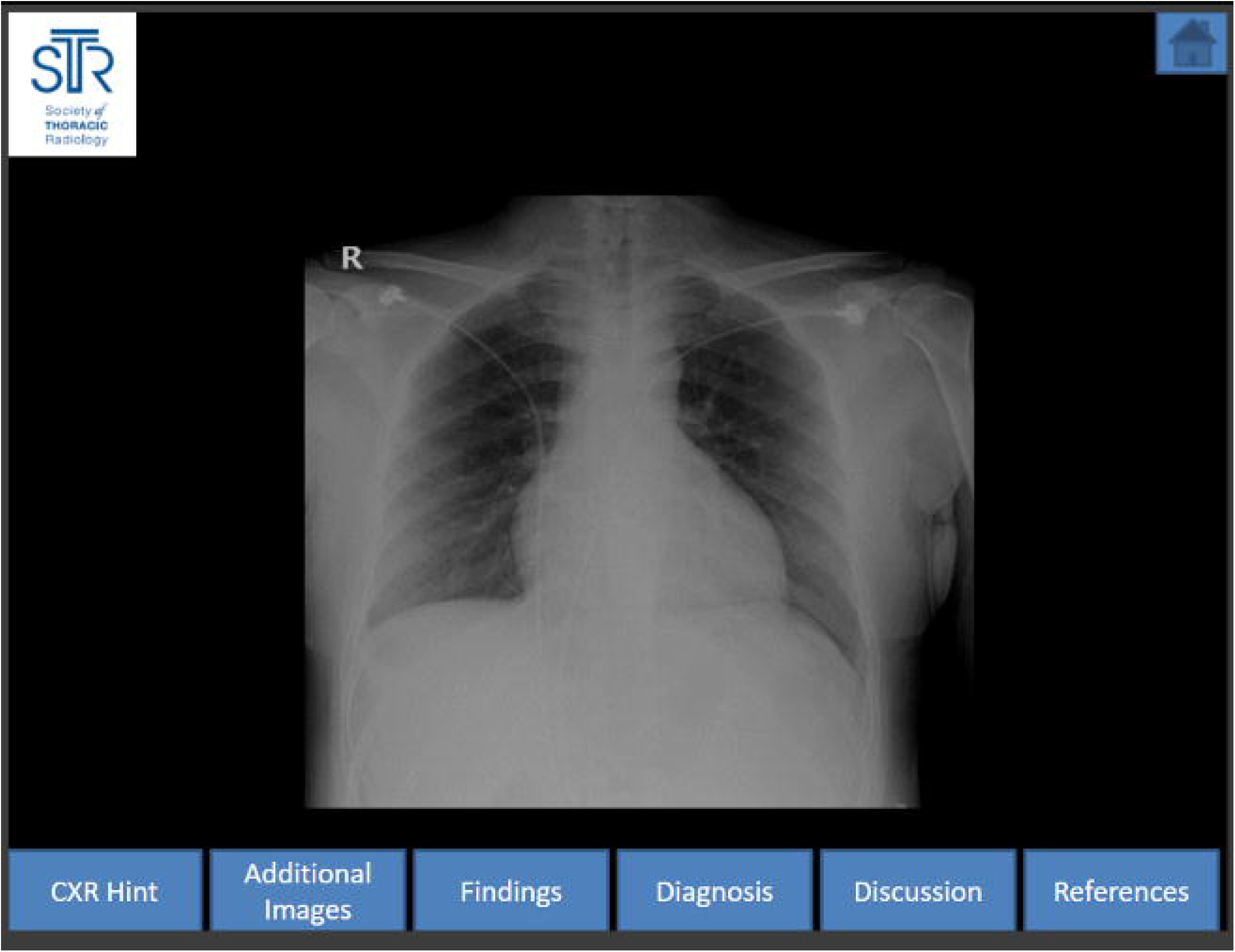
Layout of the homepage for a typical case and its sections.

### Category Classification

Cases were categorized by the cardiac radiologists into five broad groups based on cardiac categories: cardiomyopathies (CMP) (n=20), valvular/vascular pathologies (n= 17), congenital heart diseases (CHD) (n=15), and coronary artery diseases (CAD) (n=15), neoplasms (n=13). The diagnostic performances of LLMs and radiologists were compared for each group.

### Study Sessions and Performance Evaluation of the Radiologists

Three general radiologist-I (E.Ç.), Radiologist-II (T.C.), and Radiologist-III (Y.C.G.), each with six years of experience, and a cardiac radiologist (M.D.) with 15 years of experience independently assessed the cases. None of the radiologists had previously attempted to resolve the “Cardiac Case of the Month” on the Society of Thoracic Radiology website.

Each radiologist created a list of single correct answers and three possible differential diagnoses for each case.

In the first, unassisted, session, Radiologists-I, II, and the Cardiac Radiologist reviewed the patient’s medical history and visually assessed the images without seeing the text-based explanations of the images provided in the Findings section and without assessing the information in the Hint section.

Radiologist-III has solved the cases using only text-based information, the same information provided to the LLMs, instead of visual information.

Just after the first section, all radiologists re-created their diagnosis and differential diagnosis lists with the assistance of ChatGPT-4o. We chose ChatGPT-4o as the assistant LLM because it was the most up-to-date model available at the beginning of the study.

Owing to their text-based nature, the LLMs were provided with the history and findings sections to solve the cases.

The Cardiac Radiologist evaluated all diagnoses and differential diagnoses provided by the general radiologists and LLMs. All general radiologists evaluated the Cardiac Radiologist’s performance by consensus. The evaluation process was based on the correct diagnosis and differential diagnosis lists provided by the Society of Thoracic Radiology website for each case.

An overview of the workflow is shown in Figure 2.

**Figure 2.**
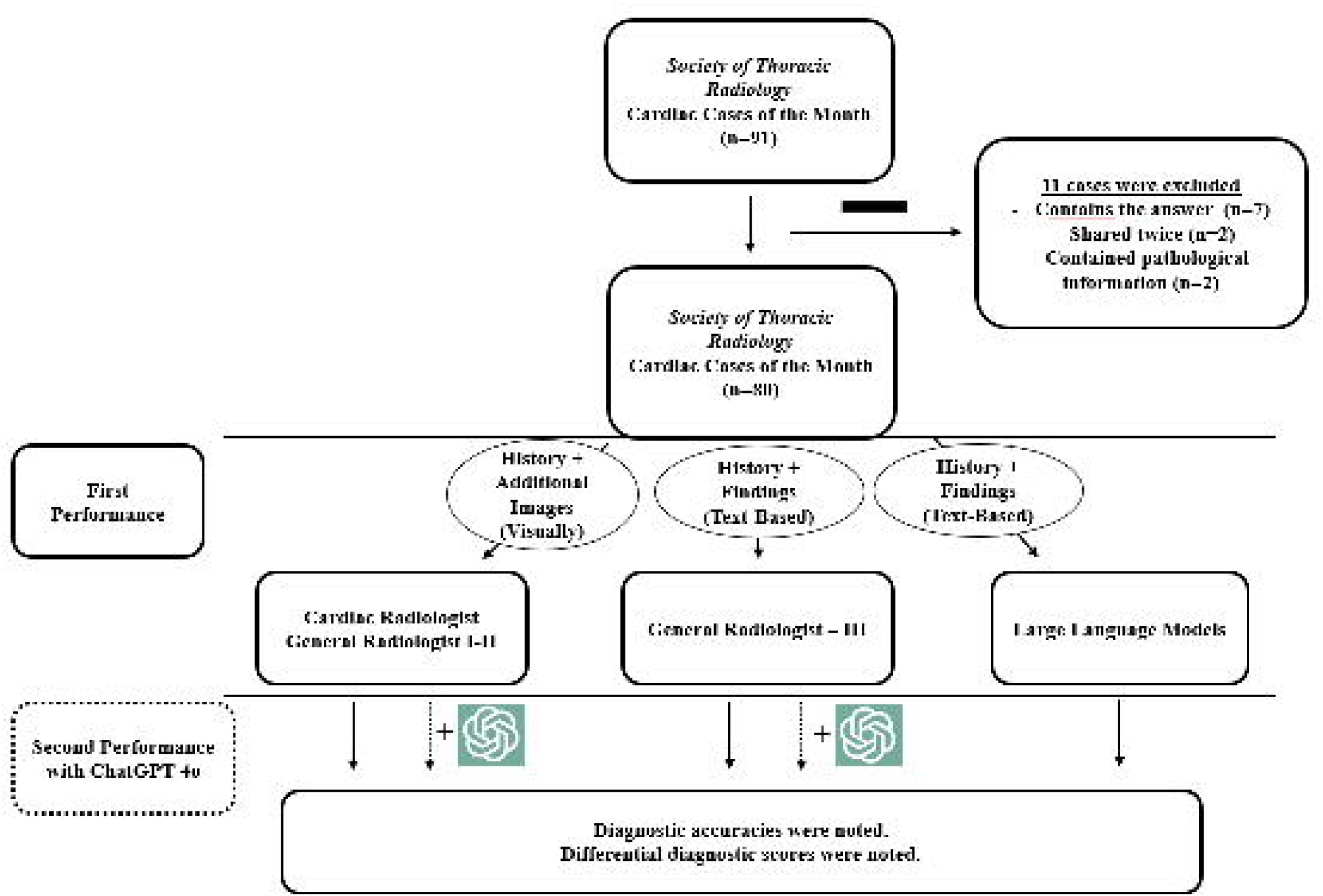
Workflow of the study.

### Prompt Design for LLMs

To achieve the best possible performance from each model, we used a similar prompt that had demonstrated superior results in a previous study which evaluates the diagnostic performance of ChatGPT 3.5 and 4 in thoracic radiology cases (15). The input prompt used in our study was: “As a highly experienced Professor of Radiology with 30 years of expertise in cardiac imaging, you assist in solving cardiac radiology cases. Your task is to analyze patient histories and imaging findings to determine the most likely diagnosis, as well as provide three differential diagnoses for each case below. To complete this task, review the patient history and imaging findings provided for each case, analyze the data thoroughly, utilize your extensive knowledge in cardiac imaging, ensure that your diagnoses are well-supported, and make thoughtful decisions.”

This prompt was presented in May 2024 on eleven different LLMs with default hyperparameters: OpenAI’s ChatGPT-4o, 4, and 3.5 (https://chat.openai.com), Google Gemini 1.5 Flash, 1.5 Pro, and 1.0 (https://gemini.google.com/), Perplexity (https://perplexity.ai), Claude 3 Opus, 3.5 Sonnet and 3 Sonnet (https://claude.ai), Mistral Large (https://mistral.ai), and Meta 3 Llama 70b (https://llama.meta.com/llama3). Patient histories and imaging findings were added sequentially as text-based information to the same chat session for each LLM, and responses were recorded.

It’s important to mention that the LLMs in this study were not pre-trained with specific commands or questions. Each inquiry was posed within a single chat session, without initiating new chat tabs for separate questions.

In the ChatGPT-4o assisted second performance of the Cardiac Radiologist and Radiologist-I and II, the same prompt was used, incorporating the patient’s history and the findings they generated after their visual assessments. Similarly, Radiologist-III generated additional findings based on the findings of the cases and used the same prompt to receive ChatGPT-4o assistance.

### Differential Diagnosis Score (DDx Score)

The diagnosis section of the cases included the differential diagnoses for each case. Using this information as the gold standard, both radiologists and LLMs were scored between 1 and 5 for differential diagnosis accuracy. The differential diagnosis scores (DDx Score) for LLMs and General Radiologists were assigned by the Cardiac Radiologist. The Cardiac Radiologist’s DDx score was determined by consensus among the general radiologists, ensuring that no one knew their score to prevent potential bias.

Scoring for the differential diagnosis list was performed as follows:

- Five points (excellent): both diagnosis and differential diagnosis were correct.
- Four points (good): when the diagnosis is correct, the differential diagnosis is incomplete.
- Three points (moderate): when the diagnosis was incorrect, the correct diagnosis was at the top of the differential list.
- Two points (poor): when the diagnosis was incorrect but the correct diagnosis was at the bottom of the differential list.
- One point (very poor): when both diagnosis and differential diagnosis were irrelevant.

### Performance Evaluation of LLMs

In separate sessions, demographic information and medical histories of each case from the History section of the Society of Thoracic Radiology website, as well as the text-based information in the Findings section, were provided to LLMs. Information in the Hint section has been omitted to prevent bias.

As all LLMs do not have multimodal capabilities, the images were not visually evaluated. The LLMs were tasked with providing a correct answer and a list of three differential diagnoses for each case, based on the provided text-based information. The accuracy of the responses and differential diagnosis lists provided by the LLMs were independently assessed by a Cardiac Radiologist (M.D.) using the DDx Score criteria mentioned above.

### Statistical Analysis

We used descriptive statistics, including means, standard deviations, medians, minima, maxima, and percentages. The distribution of variables was checked using the Kolmogorov-Smirnov test. For comparing quantitative data, we applied the Kruskal-Wallis test, followed by Tamhane’s T2 test for multiple post-hoc comparisons. We used the chi-square test to compare the qualitative data. We used McNemar’s test to compare diagnostic accuracy, and the Wilcoxon test was used to compare the DDx Scores of both LLMs and radiologists. All statistical analyses were performed using SPSS 28.0, with significance set at P<0.05.

## RESULTS

### Diagnostic Accuracy of LLMs and Radiologists

A total of 80 “Cardiac Case of the Month” were included in this study. Initially, the diagnostic accuracy for the Cardiac Radiologist was 72.5% (n=58), for General Radiologist-I was 53.8% (n=43), and for General Radiologist-II was 51.3% (n=41), who evaluated cases visually (Table 1) (Figure 3).

**Figure 3.**
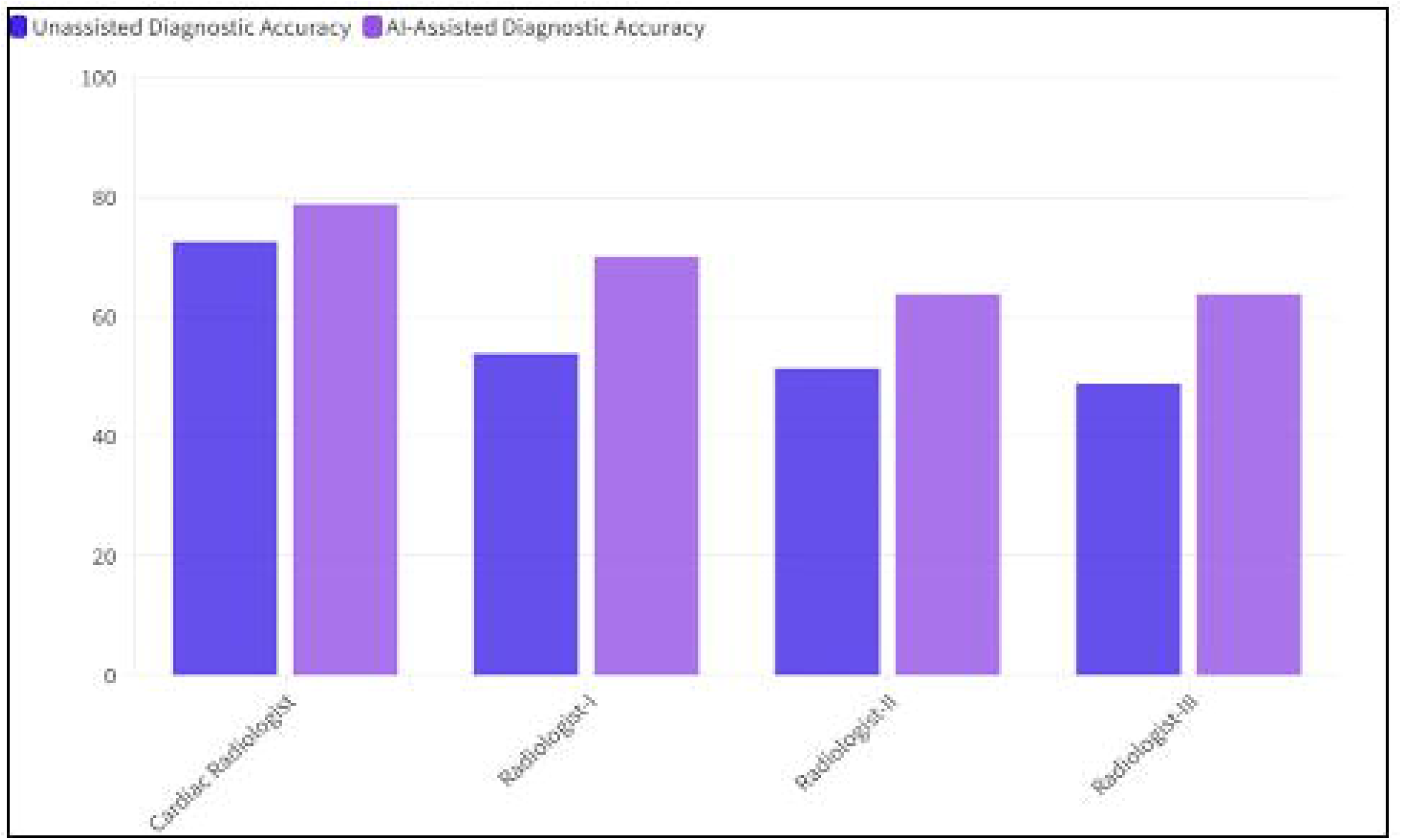
Diagnostic accuracy percentages of the radiologists with and without the assistance of ChatGPT-4o for Cardiac Cases of Month.

**Table 1.**
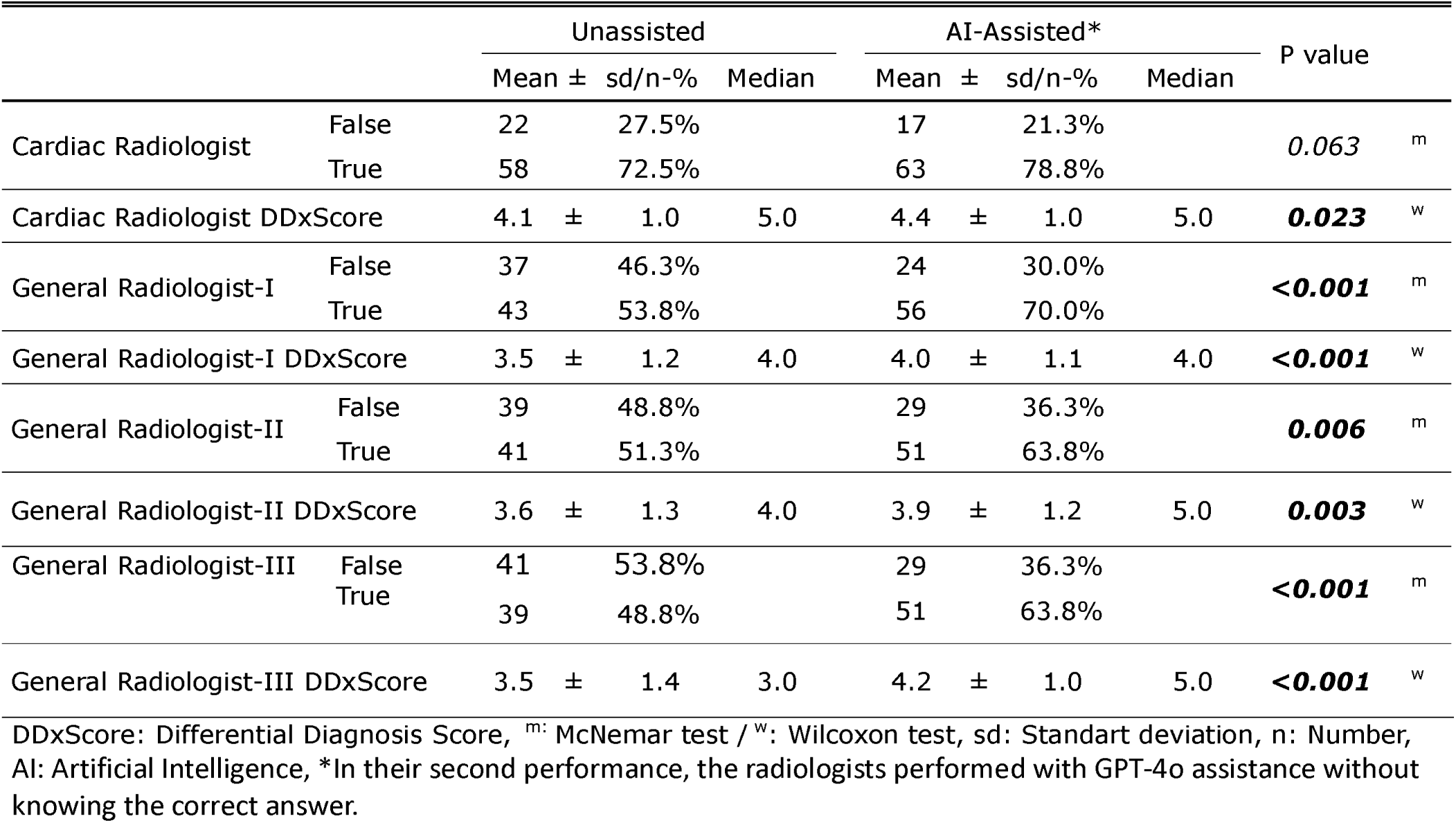
Diag nostic performance of radiolog ists.

With the assistance of ChatGPT-4o, the diagnostic accuracies were improved to 78.8% (n=63) for the Cardiac Radiologist, 70.0% (n=56) for General Radiologist-I, and 63.8% (n=51) for General Radiologist-II (Table 1). The improvements in diagnostic accuracy for both General Radiologist-I and General Radiologist-II were statistically significant (P≤0.006), whereas the improvement for the Cardiac Radiologist was not (P=0.063) (Table 1).

With the assistance of ChatGPT-4o, the diagnostic accuracies of both General Radiologist-I and II showed no significant difference than the unassisted performance of Cardiac Radiologist (P>0.05) (Table 2).

**Table 2.** Comparison of diagnostic accuracy of radiologists who solved cases with visual information: P-values obtained from.

|  | **AI-Assisted**<br>**Cardiac Radiologist** | **Cardiac Radiologist** | **AI-Assisted**<br>**Radiologist-I** | **Radiologist-I** | **AI-Assisted**<br>**Radiologist-II** | **Radiologist-II** |
| --- | --- | --- | --- | --- | --- | --- |
| **AI-Assisted**<br>**Cardiac**<br>**Radiologist** | - | 0.063 | 0.143 | <0.001 | 0.012 | <0.001 |
| **Cardiac**<br>**Radiologist** | 0.063 | - | 0.804 | 0.003 | 0.143 | <0.001 |
| **AI-Assisted**<br>**Radiologist-**<br>**I** | 0.143 | 0.804 | - | <0.000 | 0.359 | 0.003 |
| **Radiologist-**<br>**I** | <0.001 | 0.003 | <0.000 | - | 0.077 | 0.815 |
| **AI-Assisted**<br>**Radiologist-**<br>**II** | 0.012 | 0.143 | 0.359 | 0.077 | - | 0.006 |
| **Radiologist-**<br>**II** | <0.001 | <0.001 | 0.003 | 0.815 | 0.006 | - |

Among the LLMs and General Radiologist-III, who assessed cases based on text-based information, Claude 3 Opus and Claude 3.5 Sonnet demonstrated the highest performance, correctly answering 81.3% (n=65) of the cases. This was followed by Claude 3 Sonnet with 70.0% (n=56), ChatGPT-4o with 67.5% (n=54), Mistral Large with 65.0% (n=52), ChatGPT-4 with 63.8% (n=51), ChatGPT-3.5 and Google Gemini 1.5 Pro with 62.5% (n=50), Google Gemini 1.5 Flash and Meta Llama 3 70b with 60.0% (n=48), Perplexity with 58.8% (n=47), and Google Gemini 1.0 with 56.3% (n=45) (Table 3) (Figure 4).

**Figure 4.**
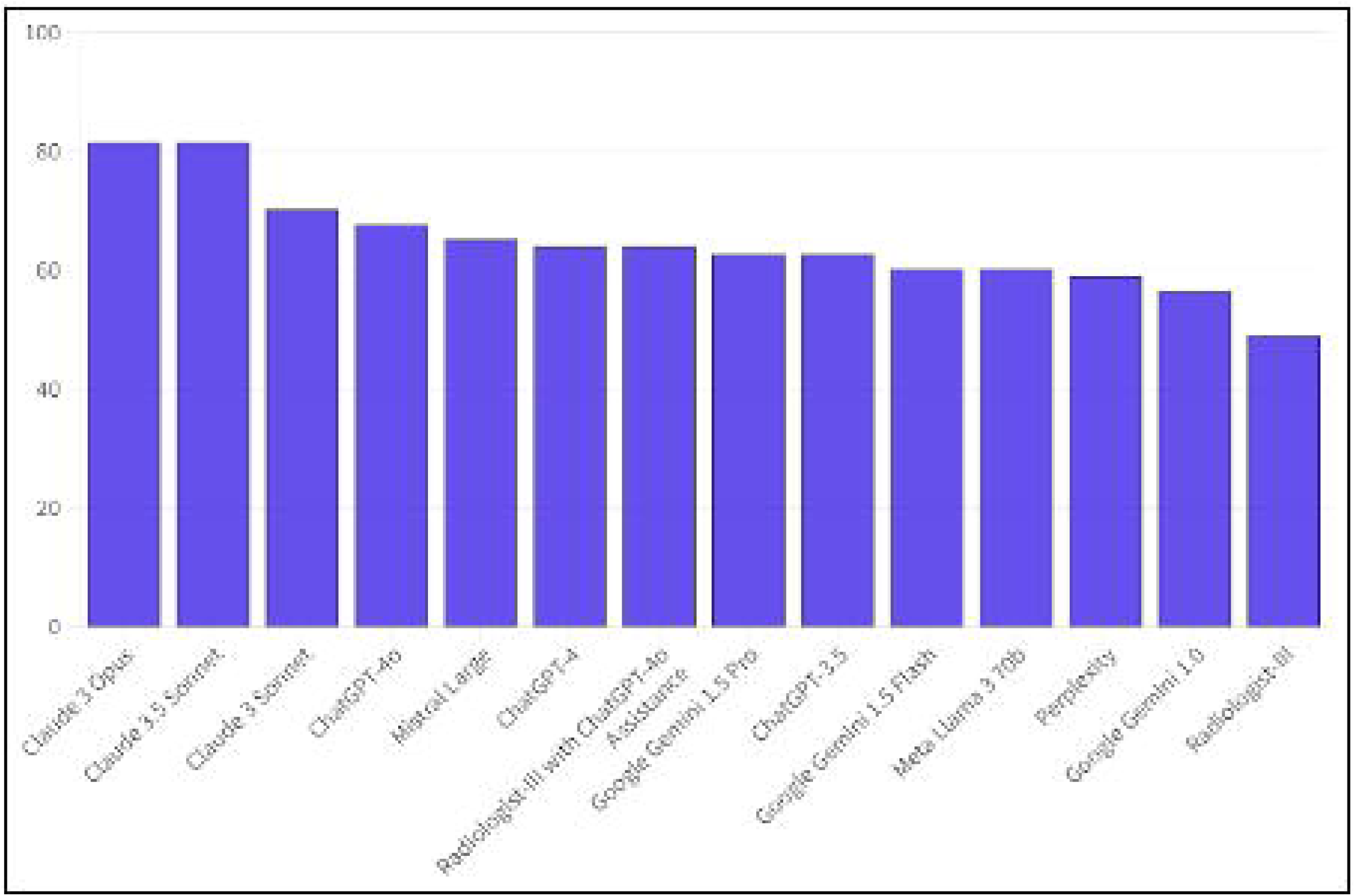
Diagnostic accuracy percentages of the large language models and Radiologist-III, who solved cases with-text based information for Cardiac Cases of Month.

**Table 3.**
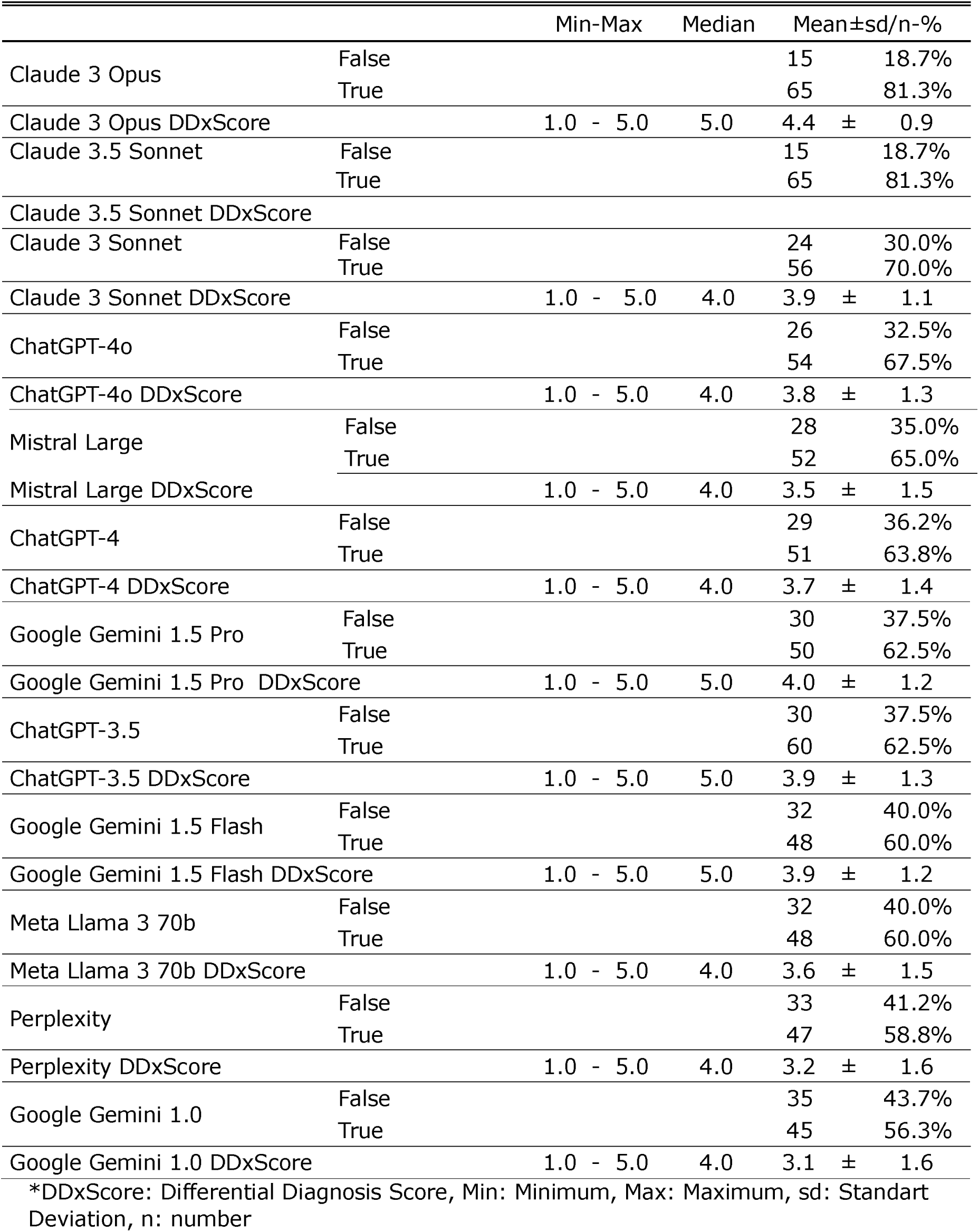
Diagnostic performance of large language models.

|  |  | Min-Max | Median | Mean±sd/n-% |  |
| --- | --- | --- | --- | --- | --- |
| Claude 3 Opus | False |  |  | 15 | 18.7% |
|  | True |  |  | 65 | 81.3% |
| Claude 3 Opus DDxScore |  | 1.0 - 5.0 | 5.0 | 4.4 ± 0.9 |  |
| Claude 3.5 Sonnet | False |  |  | 15 | 18.7% |
|  | True |  |  | 65 | 81.3% |
| Claude 3.5 Sonnet DDxScore |  |  |  |  |  |
| Claude 3 Sonnet | False |  |  | 24 | 30.0% |
|  | True |  |  | 56 | 70.0% |
| Claude 3 Sonnet DDxScore |  | 1.0 - 5.0 | 4.0 | 3.9 ± 1.1 |  |
| ChatGPT-4o | False |  |  | 26 | 32.5% |
|  | True |  |  | 54 | 67.5% |
| ChatGPT-4o DDxScore |  | 1.0 - 5.0 | 4.0 | 3.8 ± 1.3 |  |
| Mistral Large | False |  |  | 28 | 35.0% |
|  | True |  |  | 52 | 65.0% |
| Mistral Large DDxScore |  | 1.0 - 5.0 | 4.0 | 3.5 ± 1.5 |  |
| ChatGPT-4 | False |  |  | 29 | 36.2% |
|  | True |  |  | 51 | 63.8% |
| ChatGPT-4 DDxScore |  | 1.0 - 5.0 | 4.0 | 3.7 ± 1.4 |  |
| Google Gemini 1.5 Pro | False |  |  | 30 | 37.5% |
|  | True |  |  | 50 | 62.5% |
| Google Gemini 1.5 Pro DDxScore |  | 1.0 - 5.0 | 5.0 | 4.0 ± 1.2 |  |
| ChatGPT-3.5 | False |  |  | 30 | 37.5% |
|  | True |  |  | 60 | 62.5% |
| ChatGPT-3.5 DDxScore |  | 1.0 - 5.0 | 5.0 | 3.9 ± 1.3 |  |
| Google Gemini 1.5 Flash | False |  |  | 32 | 40.0% |
|  | True |  |  | 48 | 60.0% |
| Google Gemini 1.5 Flash DDxScore |  | 1.0 - 5.0 | 5.0 | 3.9 ± 1.2 |  |
| Meta Llama 3 70b | False |  |  | 32 | 40.0% |
|  | True |  |  | 48 | 60.0% |
| Meta Llama 3 70b DDxScore |  | 1.0 - 5.0 | 4.0 | 3.6 ± 1.5 |  |
| Perplexity | False |  |  | 33 | 41.2% |
|  | True |  |  | 47 | 58.8% |
| Perplexity DDxScore |  | 1.0 - 5.0 | 4.0 | 3.2 ± 1.6 |  |
| Google Gemini 1.0 | False |  |  | 35 | 43.7% |
|  | True |  |  | 45 | 56.3% |
| Google Gemini 1.0 DDxScore |  | 1.0 - 5.0 | 4.0 | 3.1 ± 1.6 |  |

\*DDxScore: Differential Diagnosis Score, Min: Minimum, Max: Maximum, sd: Standard Deviation, n: number

The diagnostic accuracy of General Radiologist-III was 48.8% (n=39), which improved to 63.8% (n=51) with the assistance of ChatGPT-4o, a statistically significant increase (P<0.001) (Table 4).

**Table 4.** Comparison of diagnostic accuracy of LLMs and Radiologist-III who solved cases with text-based. information: P-values obtained from the McNemar test

|  | Claude<br>3 Opus | Claude<br>3.5<br>Sonnet | Claude<br>3<br>Sonnet | Chat<br>GPT-4o | Mistral<br>Large | Chat<br>GPT-4 | Gemini<br>1.5 Pro | Chat<br>GPT-3.5 | Gemini<br>1.5<br>Flash | Llama<br>3 70b | Perple<br>xity | AI-Assisted<br>Radiologist-III | Radiologist-III |
| --- | --- | --- | --- | --- | --- | --- | --- | --- | --- | --- | --- | --- | --- |
| Claude 3<br>Opus | - | 1 | 0.049 | 0.013 | 0.011 | 0.003 | 0.001 | 0.001 | 0.003 | <0.001 | <0.001 | 0.003 | <0.001 |
| Claude<br>3.5<br>Sonnet | 1 | - | 0.035 | 0.013 | 0.007 | 0.003 | 0.001 | 0.003 | 0.002 | <0.001 | <0.001 | 0.004 | <0.001 |
| Claude 3<br>Sonnet | 0.049 | 0.035 | - | 0.815 | 0.503 | 0.332 | 0.263 | 0.286 | 0.115 | 0.057 | 0.078 | 0.383 | 0.002 |
| Chat<br>GPT-4o | 0.013 | 0.013 | 0.815 | - | 0.804 | 0.664 | 0.454 | 0.571 | 0.327 | 0.210 | 0.189 | 0.664 | 0.003 |
| Mistral<br>Large | 0.011 | 0.007 | 0.503 | 0.804 | - | 1 | 0.815 | 0.832 | 0.454 | 0.454 | 0.302 | 1 | 0.011 |
| Chat<br>GPT-4 | 0.003 | 0.003 | 0.332 | 0.664 | 1 | - | 1 | 1 | 0.648 | 0.581 | 0.454 | 1 | 0.017 |
| Gemini<br>1.5 Pro | 0.001 | 0.001 | 0.263 | 0.454 | 0.815 | 1 | - | 1 | 0.804 | 0.815 | 0.648 | 1 | 0.013 |
| Chat<br>GPT-3.5 | 0.001 | 0.003 | 0.286 | 0.571 | 0.832 | 1 | 1 | - | 0.804 | 0.832 | 0.664 | 1 | 0.027 |
| Gemini<br>1.5 Flash | 0.003 | 0.002 | 0.115 | 0.327 | 0.454 | 0.648 | 0.804 | 0.804 | - | 1 | 1 | 0.678 | 0.064 |
| Llama<br>3 70b | <0.001 | <0.001 | 0.057 | 0.210 | 0.454 | 0.581 | 0.815 | 0.832 | 1 | - | 1 | 0.664 | 0.093 |
| Perplexit<br>y | <0.001 | <0.001 | 0.078 | 0.189 | 0.302 | 0.454 | 0.648 | 0.664 | 1 | 1 | - | 0.503 | 0.152 |
| AI-Assisted<br>Radiologi<br>st-III | 0.003 | 0.004 | 0.383 | 0.664 | 1 | 1 | 1 | 1 | 0.678 | 0.664 | 0.503 | - | <0.001 |
| Radiologi<br>st-III | <0.001 | <0.001 | 0.002 | 0.003 | 0.011 | 0.017 | 0.013 | 0.027 | 0.064 | 0.093 | 0.152 | <0.001 | - |

<sup>2</sup>perplexity's information was not given because it did not fit into the table structure that could fit on a single page.

Claude 3 Opus’s and Claude 3.5 Sonnet’s diagnostic accuracy was significantly superior to all other LLMs and performances of Radiologist-III (P<0.05) (Table 4). Claude 3 Sonnet’s accuracy was significantly higher than that of Google Gemini 1.0 (P=0.019) and Radiologist-III’s unassisted performance (P=0.002), although it was not significantly different from other LLMs or Radiologist-3’s second performance (P>0.05) (Table 4).

General Radiologist-III’s unassisted diagnostic accuracy was significantly lower than that of Claude 3 Opus, Claude 3.5 Sonnet, Claude 3 Sonnet, ChatGPT-4o, Mistral Large, ChatGPT-4, Gemini 1.5 Pro and ChatGPT-3.5 (P<0.05). Although lower than Meta Llama 3 70b, Google Gemini 1.5 Flash, Perplexity, and Google Gemini 1.0, the differences were not statistically significant (P>0.05).

### Differential Diagnosis Scores of LLMs and Radiologists

The mean DDx Score of Claude 3 Opus (4.4 ± 0.9) was significantly higher than that of all other LLMs and General Radiologist-III (P<0.05) (Table 3) (Figure 5) (see Table, Supplemental Digital Content 1, which shows the p values obtained from the Wilcoxon tests that compare DDx Scores of the LLMs and Radiologist-III).

**Figure 5.**
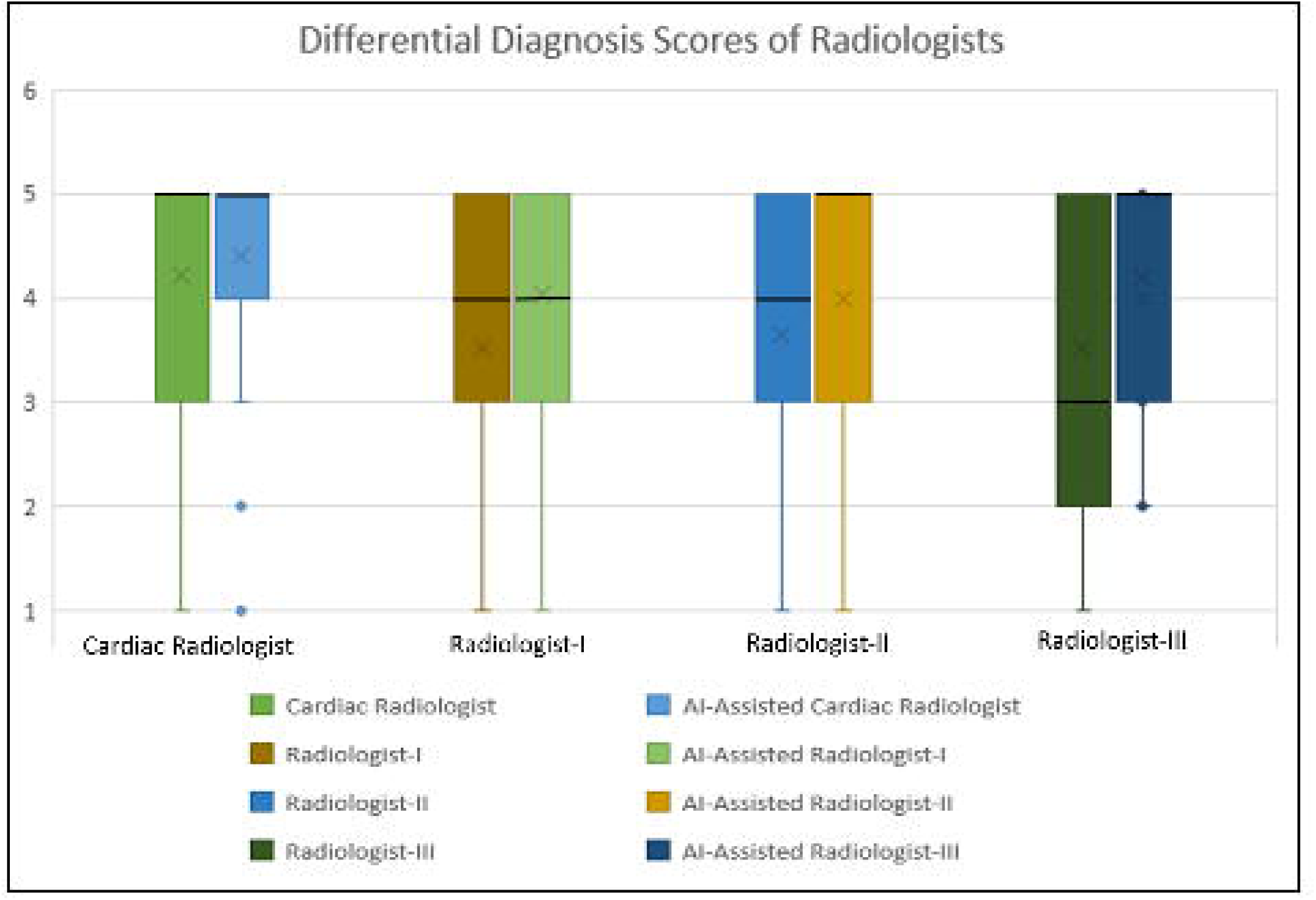
Differential Diagnosis Scores of the Large Language Models and Radiologist-III who solved cases with text-information on Cardiac Cases of Month. x: mode; black line: median.

The highest mean DDx Score among the radiologists’ unassisted performances was observed in the Cardiac Radiologist (4.2 ± 1.0), followed by Radiologist-II (3.6 ± 1.4) and General Radiologist-I (3.5 ± 1.2) (Table 1) (Figure 6). The Cardiac Radiologist’s unassisted DDx score was significantly higher than the unassisted DDx scores of both General Radiologist-I and II (P<0.001) (see Table, Supplemental Digital Content 2, which shows the p values obtained from the Wilcoxon tests that compare DDx Scores of the radiologists).

**Figure 6.**
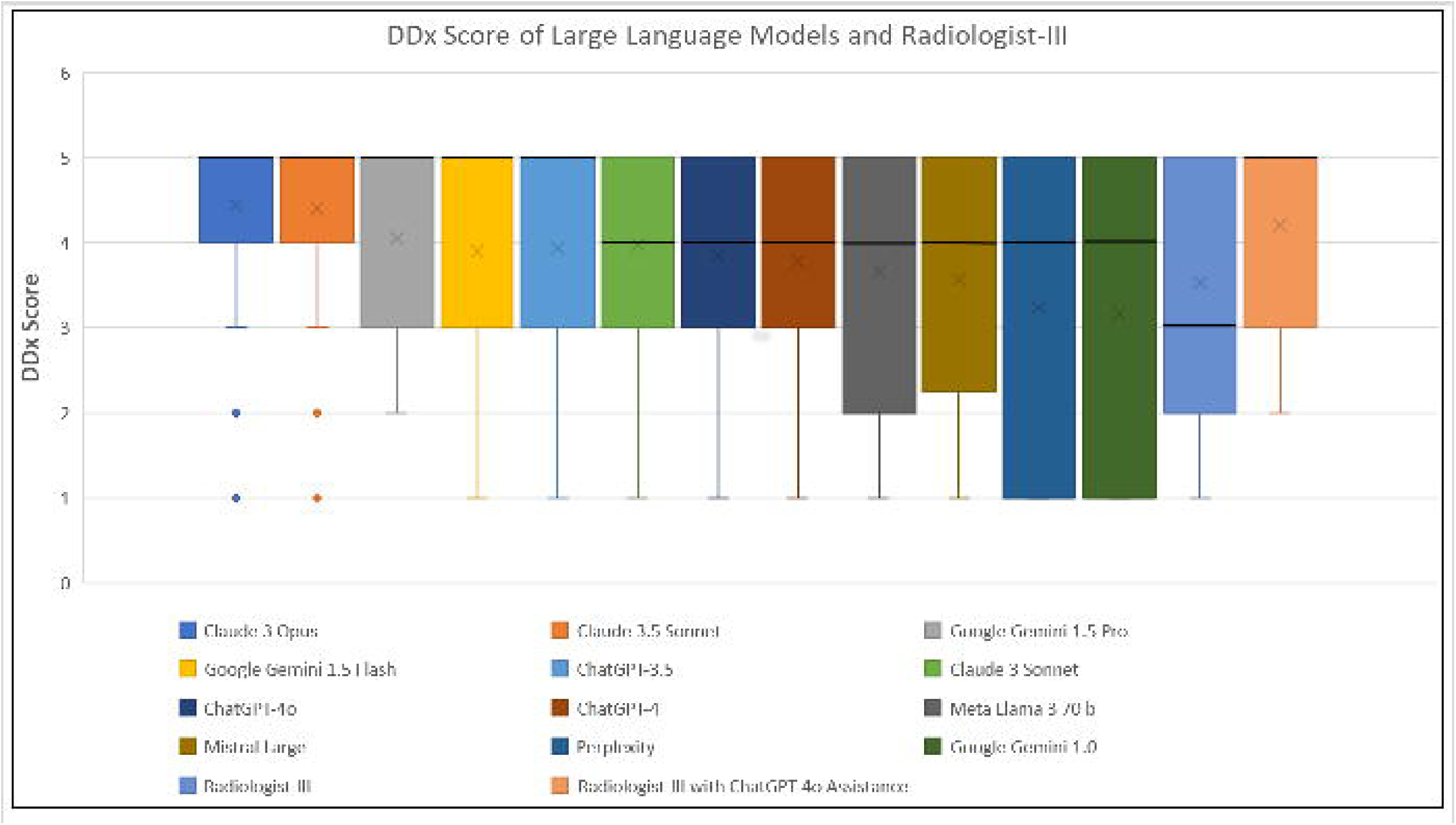
Differential Diagnosis Scores of the Radiologists with and without the assistance of ChatGPT-4o for Cardiac Cases of Month. x: mode; black line: median.

In the second performance, with the assistance of ChatGPT-4o, the Cardiac Radiologist again had the highest mean DDx score (4.4 ± 1.0), followed by Radiologist-I (4.0 ± 1.1) and Radiologist-II (3.9 ± 1.2) (Table 1). The Cardiac Radiologist’s GPT-4o assisted DDx score was significantly higher than that of both general radiologists (P≤0.004).

Remarkably, Radiologist-I’s performance with GPT-4o assistance revealed no significant difference when compared to the Cardiac Radiologist’s unassisted DDx score (P=0.151). However, the distinction between Radiologist-II’s GPT-4o assisted score and the Cardiac Radiologist’s unassisted performance remained significant (P=0.031).

All radiologists showed significant increases in their GPT-4o assisted DDx scores compared with their unassisted DDx scores (P≤0.05).

### Diagnostic Performances of LLMs and Radiologists in Categories

The accuracy rate of Google Gemini 1.5 Flash in the Neoplasms category (23.1%, n=3/13) was significantly lower than that in other categories (Chi Square test P= 0.017, adjusted Bonferroni P=0.002) (see Table, Supplemental Digital Content 3, which demonstrates performances of the LLMs across different categories).

No significant differences were observed in the accuracy rates among other LLMs and radiologists across different categories (P>0.05) (see Tables, Supplemental Digital Content 3 and 4, which demonstrate performances of the LLMs and radiologists across different categories).

When comparing the DDx Score across categories, a statistically significant difference was found between the DDx Score of Google Gemini 1.5 Flash in the Congenital Heart Diseases category (mean: 4.46), and in the neoplasms category (mean: 3.00) (Kruskal Wallis test P= 0.036, Tamhane’s post hoc P=0.021) (see Table, Supplemental Digital Content 4, which demonstrates performances of the LLMs across different categories).

No significant group differences were detected in the DDx Scores of other LLMs and radiologists across categories (see Tables, Supplemental Digital Content 3 and 4, which demonstrate the performances of the LLMs and radiologists across different categories).

## DISCUSSION

Our study is the first to demonstrate that ChatGPT-4o may significantly improve the diagnostic performance of general radiologists for cardiac imaging. Additionally, it stands out in the literature by evaluating the performance of 12 different large language models (LLMs) in 80 specialized cardiac radiology cases.

With the assistance of ChatGPT-4o, the diagnostic accuracies of the general radiologists significantly improved (from 53.8% to 70% for General Radiologist-I and from 51.3% to 63.8% for General Radiologist-II) (P≤0.006). This improvement was not statistically significant for the Cardiac Radiologist (from 72.5% to 78.8%) (P=0.063). With ChatGPT-4o assistance, the General Radiologists’ diagnostic accuracy showed no significant difference compared to the unassisted performance of the Cardiac Radiologist (P>0.05). In the second assessment, radiologists provided ChatGPT-4o both the case history and their own findings. ChatGPT-4o then answered a diagnosis and three differential diagnoses, likely enhancing the radiologists’ knowledge or reminding them of overlooked possibilities. This led to a significant increase in diagnostic accuracy, especially for general radiologists. Our methodology, which used a single prompt for each case, was designed to show the minimum potential contribution of LLMs as radiological assistants. With tailored and an unlimited number of prompts addressing specific case details, LLMs could achieve even more successful outcomes by providing highly precise differential diagnoses (2,15).

Regarding Differential Diagnosis Score (DDx Score), all radiologists showed significant increases in their GPT-4o assisted DDx Scores compared to their unassisted DDx Scores (P<0.05). Remarkably, Radiologist-I’s DDx Score with GPT-4o assistance (4.0 ± 1.1) revealed no significant difference when compared to the Cardiac Radiologist’s unassisted DDx score (4.2 ± 1.0) (P=0.151). However, the distinction between Radiologist-II’s GPT-4o assisted score (3.9 ± 1.2) and the Cardiac Radiologist’s unassisted performance remained significant (P=0.031). These findings highlight the potential of LLMs, such as ChatGPT-4o, in helping general radiologists generate more comprehensive and accurate differential diagnoses, thus narrowing the performance gap between them and specialized cardiac radiologists.

In a study similar to ours, Siepmann et al. assessed the diagnostic performance of six radiologists with different levels of expertise on a set of 40 radiographs focused on general radiology (20). The radiologists evaluated the cases both unassisted and with the assistance of the ChatGPT-4, and their confidence in their diagnoses was measured after each assessment. The findings revealed that the ChatGPT-4-assisted diagnoses were marginally more accurate and considerably more confident than the unassisted diagnoses. However, researchers also observed that ChatGPT-4 provided potentially harmful information in 7.4% of cases. Siepmann et al.’s findings aligned with ours, revealing that less experienced radiologists exhibited a more substantial enhancement in their diagnostic accuracy when using ChatGPT-4 as an assistive tool, in comparison to the improvements observed among their more seasoned colleagues (20).

The best-performing LLMs, Claude 3 Opus and Claude 3.5 Sonnet achieved a diagnostic accuracy of 81.3%, surpassing the accuracy of all other LLMs, followed by Claude 3 Sonnet (70.0%) and ChatGPT-4o (67.5%). Regarding the differential diagnosis score (DDx Score), the DDx Score of Claude 3 Opus (4.4 ± 0.9) and Claude 3.5 Sonnet (4.4±1.0) was significantly higher than that of all other LLMs, too. The potential reasons behind Claude 3 Opus’s and Claude 3.5 Sonnet’s superior performance compared to other models may be related to factors such as the model’s architecture, training data, or the fine-tuning process. Understanding the key drivers of its success could inform the development and refinement of future AI tools in radiology.

Sonoda et al. tested the three LLMs on “Radiology Diagnosis Please” cases (16). They also found a ranking among LLMs similar to ours, with Claude 3 Opus demonstrating a diagnostic accuracy of 54%, GPT-4o 41%, and Gemini 1.5 Pro 33.9% (16).

Gunes et al. measured the knowledge of LLMs regarding Middle Meningeal Artery Embolization by presenting 20 questions on this topic (22). The results were compelling: Meta Llama3 70b achieved the highest accuracy at 95% (19/20 questions), followed by Claude Opus, ChatGPT 4, and ChatGPT 3.5, each with an accuracy of 90% (18/20 questions) (22).

In a study of 42 publicly available questions related to radiation protection and safety, Camur et al. found that Claude 3 Opus and ChatGPT 4 Turbo achieved the highest accuracy of 87.1% (34/39 questions). They were followed by Claude Sonnet at 79.4% (31/39 questions), and Google Gemini 1.5 Pro 1M and Meta Llama 3 70B, each at 76.9% (30/39 questions) (23).

The varying rankings of LLMs across studies can be attributed to several factors. Differences in the training data can result in some models performing better in specific fields, especially if they are trained on larger datasets related to that area. The difficulty level of the questions also plays a role; some models may excel in answering more complex or nuanced queries, whereas others may struggle. Additionally, the sample size can impact the results; a smaller sample size may not accurately represent the models’ true capabilities.

In our study, General Radiologist-III solved the cases using the same text-based information provided to the LLMs, achieving a diagnostic accuracy of 48.8% on his first performance. This accuracy was significantly lower than that of Claude 3 Opus and Claude 3.5 Sonnet (81.3%), Claude 3 Sonnet (70.0%), ChatGPT-4o (67.5%), Mistral Large (65.0%), ChatGPT-4 (63.8%), and ChatGPT-3.5 (62.5%), and Google Gemini 1.5 Pro (62.5%) (P<0.05). While his accuracy was also lower than that of Meta LLama 3 70b (60.0%), Google Gemini 1.5 Flash (60.0%), Perplexity (58.8%), and Google Gemini 1.0 (56.3%), these differences were not statistically significant (P>0.05).

Relying purely on text-based information, most LLMs outperformed General Radiologist-III, indicating that their strong natural language understanding capabilities can partially compensate for the lack of image interpretation.

Although our study contributes significantly to the cardiac radiology knowledge of LLMs, it has several limitations. First, we used a single publicly available case series, which may not fully represent the variety and complexity of real-world cardiac radiology practices. Second, using only a single prompt might have influenced both the performance of the LLMs and radiologists’ second assessments. Different prompts could potentially yield better or worse diagnostic accuracy (2,15). Third, we evaluated the performance of only ChatGPT-4o as an assistant, even though the it was not the most successful LLM in our study. This suggests that the radiologists’ second performance might have yielded better results if Claude 3 Opus or Claude 3.5 Sonnet models had been used. Four, books or radiology-focused online references were deliberately excluded according to the study design (20). This limitation resulted in a setup that does not accurately reflect real-world radiologic practice. Future research should include comparisons between established online resources and LLMs to assess their respective values in the reading room. Finally, future studies should expand the dataset, incorporate different LLMs as AI assistants, and evaluate the performance of radiologists and whether LLMs affect their diagnostic performance. Additionally, since the datasets of these LLMs are not available beyond their training data, it is unclear whether these questions were part of the original training sets, and whether LLM memorization influenced the results. Consequently, prospective studies are necessary.

In conclusion, ChatGPT-4o may significantly enhance the diagnostic performance of general radiologists for cardiac imaging, suggesting its potential as a valuable diagnostic support tool. We also revealed the superior performance of Claude 3 Opus and Claude 3.5 Sonnet in text-based cardiac radiology cases, surpassing that of other LLMs. Further research is necessary to assess the clinical utility and integration of these LLMs.

## Data Availability

All data used in the present study are available upon reasonable request to the authors

## ACKNOWLEDGMENTS

The authors express their gratitude to Onur Boyar for conducting the statistical analysis of this manuscript. They utilized ChatGPT, a language model based on the GPT-4 architecture (OpenAI; https://chat.openai.com/) to assist with grammar and English translation revisions in June 2024. The authors take full responsibility for the content of this publication and have thoroughly reviewed and edited the manuscript, as required.

## Abbreviations

AI: Artificial Intelligence
DDx: Differential Diagnosis
LLMs: Large Language Models
MRI: Magnetic Resonance Imaging
STARD: Standards for Reporting Diagnostic Accuracy Studies

---

**Supplementary Digital Content 1.** Comparison of differential diagnosis scores of LLMs and Radiologist-III who solved cases with information: P-values obtained from the Wilcoxon test.

|  | Claude 3 Opus | Claude 3.5 Sonnet | Gemini1.5 Pro | Gemini 1.5 Flash | Chat GPT-3.5 | Claude 3 Sonnet | Chat GPT-4o | Chat GPT-4 | Llama 3 70b | Mistral Large | Perplexity | AI-Assisted Radiologist-III | Radiologist-III |
| --- | --- | --- | --- | --- | --- | --- | --- | --- | --- | --- | --- | --- | --- |
| Claude 3 Opus | - | 0.661 | 0.003 | 0.001 | 0.001 | 0.001 | <0.001 | <0.001 | <0.001 | <0.001 | <0.001 | 0.032 | <0.001 |
| Claude 3.5 Sonnet | 0.661 | - |  |  |  |  |  |  |  |  |  |  |  |
| Gemini 1.5 Pro | 0.003 | 0.004 | - | 0.305 | 0.494 | 0.516 | 0.211 | 0.033 | 0.022 | 0.003 | <0.001 | 0.338 | 0.001 |
| Gemini 1.5 Flash | 0.001 | 0.002 | 0.305 | - | 0.730 | 0.634 | 0.923 | 0.398 | 0.248 | 0.026 | <0.001 | 0.068 | 0.021 |
| Chat GPT-3.5 | 0.001 | 0.004 | 0.494 | 0.730 | - | 0.907 | 0.794 | 0.444 | 0.106 | 0.034 | 0.001 | 0.119 | 0.012 |
| Claude 3 Sonnet | 0.001 | <0.001 | 0.516 | 0.634 | 0.907 | - | 0.592 | 0.172 | 0.062 | 0.044 | <0.001 | 0.071 | 0.017 |
| Chat GPT-4o | <0.001 | <0.001 | 0.211 | 0.923 | 0.794 | 0.592 | - | 0.593 | 0.337 | 0.117 | 0.002 | 0.052 | 0.063 |
| Chat GPT-4 | <0.001 | <0.001 | 0.033 | 0.398 | 0.444 | 0.172 | 0.593 | - | 0.492 | 0.347 | 0.001 | 0.006 | 0.121 |
| Llama 3 70b | <0.001 | <0.001 | 0.022 | 0.248 | 0.106 | 0.062 | 0.337 | 0.492 | - | 0.422 | 0.015 | 0.002 | 0.405 |
| Mistral Large | <0.001 | <0.001 | 0.003 | 0.026 | 0.034 | 0.044 | 0.117 | 0.347 | 0.422 | - | 0.033 | 0.001 | 0.653 |
| Perplexity | <0.001 | <0.001 | <0.001 | <0.001 | 0.001 | <0.001 | 0.002 | 0.001 | 0.015 | 0.033 | - | <0.001 | 0.201 |
| AI-Assisted Radiologist-III | 0.032 | 0.123 | 0.338 | 0.068 | 0.119 | 0.071 | 0.052 | 0.006 | 0.002 | 0.001 | <0.001 | - | <0.001 |
| Radiologist-III | <0.001 | <0.001 | 0.001 | 0.021 | 0.012 | 0.017 | 0.063 | 0.121 | 0.405 | 0.653 | 0.201 | <0.001 | - |

\*Google Gemini 1.0's information was not given because it did not fit into the table structure that could fit on a single page.

**Supplementary Digital Content 3.**
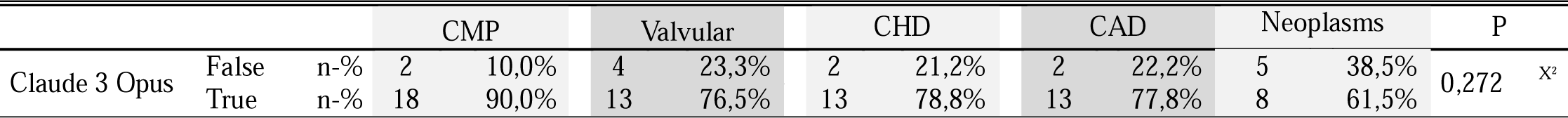
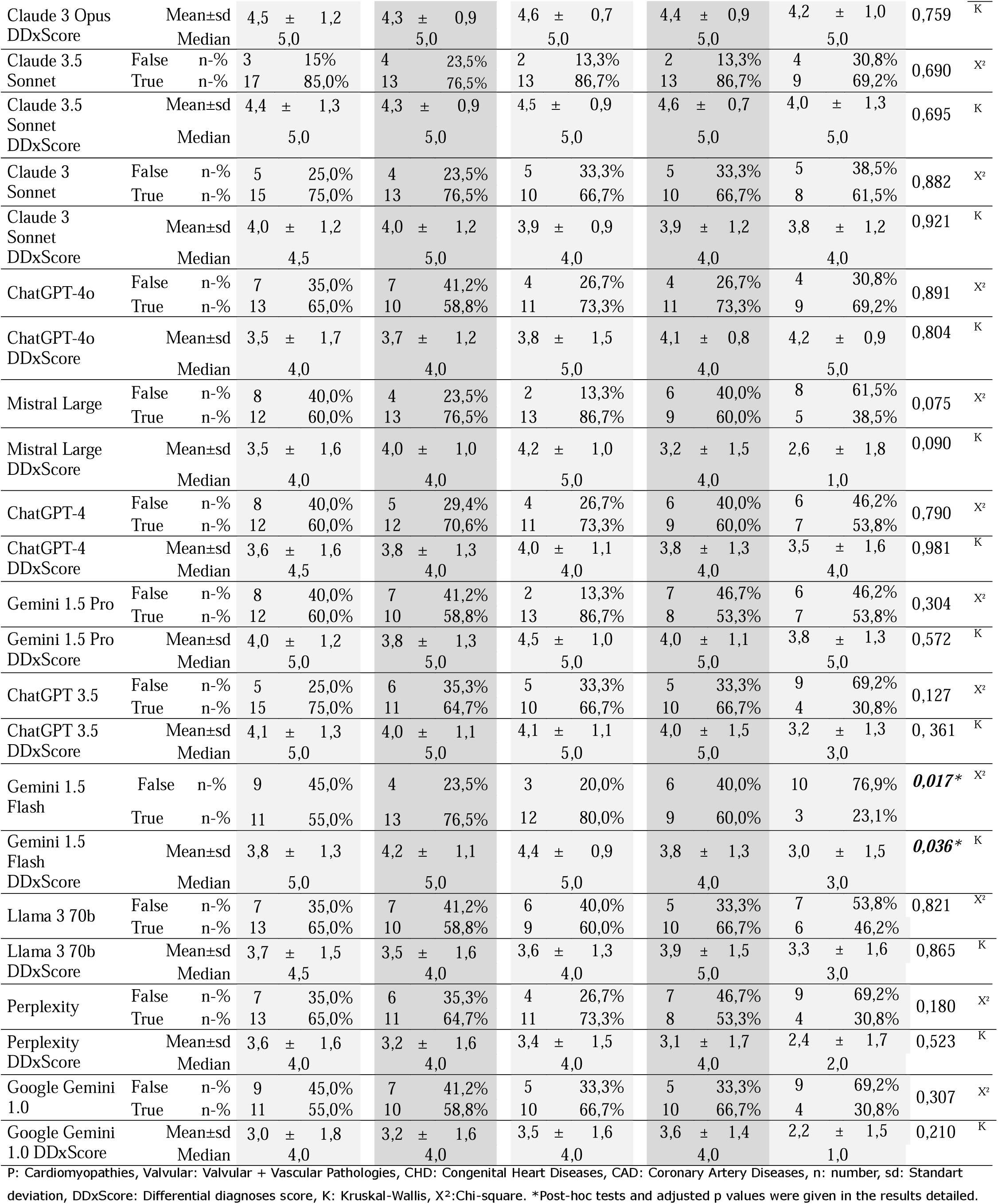
Diagnostic performance of the large language models by categories.

**Supplementary Digital Content 2.** Comparison of differential diagnosis scores of radiologists who solved cases with visual information: P-values obtained from Wilcoxon test.

|  | AI-Assisted<br>Cardiac Radiologist | Cardiac Radiologist | AI-Assisted<br>Radiologist-I | Radiologist-I | AI-Assisted<br>Radiologist-II | Radiologist-II |
| --- | --- | --- | --- | --- | --- | --- |
| **AI-Assisted**<br>**Cardiac**<br>**Radiologist** | - | 0.023 | 0.004 | <0.001 | 0.001 | <0.001 |
| **Cardiac**<br>**Radiologist** | 0.023 | - | 0.151 | <0.001 | 0.031 | <0.001 |
| **AI-Assisted**<br>**Radiologist-**<br>**I** | 0.004 | 0.151 | - | <0.001 | 0.792 | 0.006 |
| **Radiologist-**<br>**I** | <0.001 | <0.001 | <0.001 | - | <0.001 | 0.489 |
| **AI-Assisted**<br>**Radiologist-**<br>**II** | 0.001 | 0.031 | 0.792 | <0.001 | - | 0.003 |
| **Radiologist-**<br>**II** | <0.001 | <0.001 | 0.006 | 0.489 | 0.003 | - |

**Supplementary Digital Content 4.**
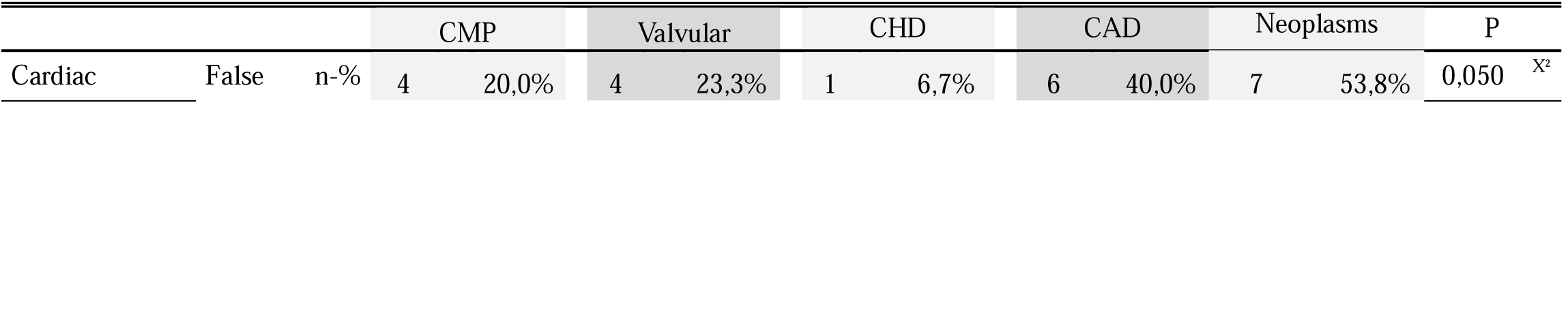
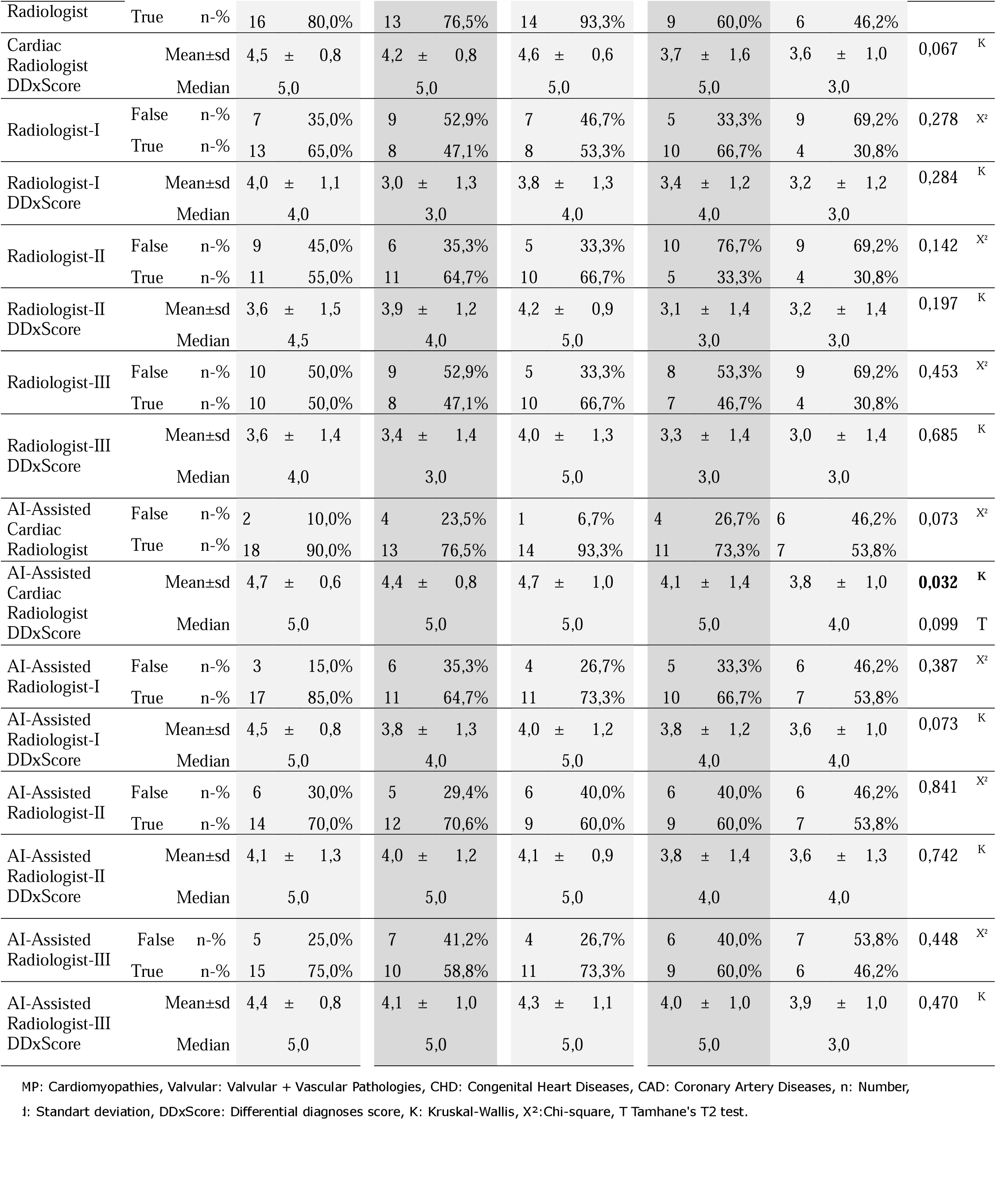
Diagnostic performance of the radiologists by categories.

|  |  |  | CMP |  | Valvular |  | CHD |  | CAD |  | Neoplasms |  | P |
| --- | --- | --- | --- | --- | --- | --- | --- | --- | --- | --- | --- | --- | --- |
| Cardiac | False | n-% | 4 | 20,0% | 4 | 23,3% | 1 | 6,7% | 6 | 40,0% | 7 | 53,8% | 0,050<sup>x2</sup> |
| Radiologist | True | n-% | 16 | 80,0% | 13 | 76,5% | 14 | 93,3% | 9 | 60,0% | 6 | 46,2% |  |
| Cardiac Radiologist | Mean±sd |  | 4,5 | ± 0,8 | 4,2 | ± 0,8 | 4,6 | ± 0,6 | 3,7 | ± 1,6 | 3,6 | ± 1,0 | 0,067<sup>K</sup> |
| DDxScore | Median |  | 5,0 |  | 5,0 |  | 5,0 |  | 5,0 |  | 3,0 |  |  |
| Radiologist-I | False | n-% | 7 | 35,0% | 9 | 52,9% | 7 | 46,7% | 5 | 33,3% | 9 | 69,2% | 0,278<sup>X²</sup> |
|  | True | n-% | 13 | 65,0% | 8 | 47,1% | 8 | 53,3% | 10 | 66,7% | 4 | 30,8% |  |
| Radiologist-I | Mean±sd |  | 4,0 | ± 1,1 | 3,0 | ± 1,3 | 3,8 | ± 1,3 | 3,4 | ± 1,2 | 3,2 | ± 1,2 | 0,284<sup>K</sup> |
|  | Median |  | 4,0 |  | 3,0 |  | 4,0 |  | 4,0 |  | 3,0 |  |  |
| Radiologist-II | False | n-% | 9 | 45,0% | 6 | 35,3% | 5 | 33,3% | 10 | 76,7% | 9 | 69,2% | 0,142<sup>X²</sup> |
|  | True | n-% | 11 | 55,0% | 11 | 64,7% | 10 | 66,7% | 5 | 33,3% | 4 | 30,8% |  |
| Radiologist-II | Mean±sd |  | 3,6 | ± 1,5 | 3,9 | ± 1,2 | 4,2 | ± 0,9 | 3,1 | ± 1,4 | 3,2 | ± 1,4 | 0,197<sup>K</sup> |
|  | Median |  | 4,5 |  | 4,0 |  | 5,0 |  | 3,0 |  | 3,0 |  |  |
| Radiologist-III | False | n-% | 10 | 50,0% | 9 | 52,9% | 5 | 33,3% | 8 | 53,3% | 9 | 69,2% | 0,453<sup>X²</sup> |
|  | True | n-% | 10 | 50,0% | 8 | 47,1% | 10 | 66,7% | 7 | 46,7% | 4 | 30,8% |  |
| Radiologist-III | Mean±sd |  | 3,6 | ± 1,4 | 3,4 | ± 1,4 | 4,0 | ± 1,3 | 3,3 | ± 1,4 | 3,0 | ± 1,4 | 0,685<sup>K</sup> |
|  | Median |  | 4,0 |  | 3,0 |  | 5,0 |  | 3,0 |  | 3,0 |  |  |
| AI-Assisted Cardiac Radiologist | False | n-% | 2 | 10,0% | 4 | 23,5% | 1 | 6,7% | 4 | 26,7% | 6 | 46,2% | 0,073<sup>X²</sup> |
|  | True | n-% | 18 | 90,0% | 13 | 76,5% | 14 | 93,3% | 11 | 73,3% | 7 | 53,8% |  |
| AI-Assisted Cardiac Radiologist | Mean±sd |  | 4,7 | ± 0,6 | 4,4 | ± 0,8 | 4,7 | ± 1,0 | 4,1 | ± 1,4 | 3,8 | ± 1,0 | **0,032**<sup>K</sup> |
|  | Median |  | 5,0 |  | 5,0 |  | 5,0 |  | 5,0 |  | 4,0 |  | 0,099<sup>T</sup> |
| AI-Assisted Radiologist-I | False | n-% | 3 | 15,0% | 6 | 35,3% | 4 | 26,7% | 5 | 33,3% | 6 | 46,2% | 0,387<sup>X²</sup> |
|  | True | n-% | 17 | 85,0% | 11 | 64,7% | 11 | 73,3% | 10 | 66,7% | 7 | 53,8% |  |
| AI-Assisted Radiologist-I | Mean±sd |  | 4,5 | ± 0,8 | 3,8 | ± 1,3 | 4,0 | ± 1,2 | 3,8 | ± 1,2 | 3,6 | ± 1,0 | 0,073<sup>K</sup> |
|  | Median |  | 5,0 |  | 4,0 |  | 5,0 |  | 4,0 |  | 4,0 |  |  |
| AI-Assisted Radiologist-II | False | n-% | 6 | 30,0% | 5 | 29,4% | 6 | 40,0% | 6 | 40,0% | 6 | 46,2% | 0,841<sup>X²</sup> |
|  | True | n-% | 14 | 70,0% | 12 | 70,6% | 9 | 60,0% | 9 | 60,0% | 7 | 53,8% |  |
| AI-Assisted Radiologist-II | Mean±sd |  | 4,1 | ± 1,3 | 4,0 | ± 1,2 | 4,1 | ± 0,9 | 3,8 | ± 1,4 | 3,6 | ± 1,3 | 0,742<sup>K</sup> |
|  | Median |  | 5,0 |  | 5,0 |  | 5,0 |  | 4,0 |  | 4,0 |  |  |
| AI-Assisted Radiologist-III | False | n-% | 5 | 25,0% | 7 | 41,2% | 4 | 26,7% | 6 | 40,0% | 7 | 53,8% | 0,448<sup>X²</sup> |
|  | True | n-% | 15 | 75,0% | 10 | 58,8% | 11 | 73,3% | 9 | 60,0% | 6 | 46,2% |  |
| AI-Assisted Radiologist-III | Mean±sd |  | 4,4 | ± 0,8 | 4,1 | ± 1,0 | 4,3 | ± 1,1 | 4,0 | ± 1,0 | 3,9 | ± 1,0 | 0,470<sup>K</sup> |
|  | Median |  | 5,0 |  | 5,0 |  | 5,0 |  | 5,0 |  | 3,0 |  |  |

MP: Cardiomyopathies, Valvular: Valvular + Vascular Pathologies, CHD: Congenital Heart Diseases, CAD: Coronary Artery Diseases, n: Number, I: Standart deviation, DDxScore: Differential diagnoses score, K: Kruskal-Wallis, X²: Chi-square, T Tamhane's T2 test.

